# Risk screening tools could potentially miss out HIV positive individuals who seek testing services: A secondary program data analysis on the performance characteristics of an adolescent and adult HIV risk screening tool in Uganda

**DOI:** 10.1101/2023.06.20.23291666

**Authors:** Marvin Lubega, Katherine Guerra, Megan Ginivan, Yewande Kamuntu, George Senyama, Andrew Musoke, Taasi Geoffrey, Sylivia Nalubega, Shaukat Khan, Matovu John Bosco Junior

**Author notes:** These authors have contributed equally to the work (Marvin Lubega, Megan Ginivan, Yewande Kamuntu, Sylivia Nalubega, Matovu John Bosco Junior). These authors also contributed equally to this work (Katherine Guerra, George Senyama, Andrew Musoke, Taasi Geoffrey, Shaukat Khan).

## Abstract

**Introduction:** Improving HIV testing efficiency has been documented to save financial and material resources for health. In October 2019, the Ministry of health Uganda deployed an HIV risk screening tool for use in 24 health facilities targeting clients aged 15 years and above in both outpatient and Voluntary Counselling and Testing departments.

**Methods:** We conducted a retrospective secondary data analysis of routinely collected HIV risk screening program data in Uganda, collected from October to November 2019, to determine the performance characteristics of the adolescent and adult HIV risk screening tool in public health facility settings. Statistical measures for the risk screening tool performance included sensitivity, specificity, positive and negative predictive values, and a cost analysis.

**Results:** A total of 19,854 clients were screened for HIV testing eligibility; we excluded 150 records with incomplete testing information. The overall positivity rate (cluster weighted prevalence of HIV) among those screened was 4.5% (95% CI: 4.1%-4.8%) versus 3.71% (95% CI: 3.06-4.50) among those not screened. The sensitivity and specificity of the risk screening tool were found to be 90.7% (95% CI: 88.4%, 92.7%) and 75.8%, (75.2-76.4) respectively. With screening, the number needed to test to identify one PLHIV reduced from 27 to 22. Although risk screening would have led to 24.5% (4,825/19,704) reduction in testing volume, 9.3% (68/732) of PLHIV would have been missed as they were misclassified as not eligible for testing. The cost per PLHIV identified fell by 3% from $69 without screening to $66.9 with implementation of the screening tool.

**Conclusions:** The use of HIV risk screening tool in OPD settings in Uganda demonstrated improved HIV testing efficiency by reducing testing volumes but resulted in screening out of a significant number of people living with HIV. The team recommends that scientifically validated HIV risk screening tools be considered for use by countries.

## Introduction

Globally, human immune-deficiency virus (HIV) testing programs have contributed to enormous progress in identifying people living with HIV (PLHIV) and increased coverage of anti-retroviral therapy (ART) (1). By 2020, Uganda was nearing the first of the United Nation’s 95-95-95 targets of identifying 95% of PLHIV with approximately 91% of the 1.4 million PLHIV already identified, 94% on treatment but with a slightly lower viral suppression rate (85%)(2). Despite remarkable progress in HIV case identification in Uganda, there were approximately 126,000 PLHIV not yet on treatment with an estimated 38,000 new HIV infections by the year 2020, indicating a need to sustain innovative HIV case identification approaches and linkage to treatment.

As Uganda gets closer to reaching identification and treatment targets, it will become increasingly difficult and resource-intensive to identify the remaining PLHIV. At the same time, resources for HIV testing services (HTS) are declining and countries are facing pressure to reduce testing volumes in order to improve efficiency by focusing on increasing HIV testing yield at different community and facility-based entry points (3). In 2017, for example, the President’s Emergency Plan for AIDS Relief (PEPFAR) had a target of conducting approximately 7.2 million HIV tests in Uganda but this target dropped to just over 1.7 million in 2020, with targets for provider-initiated testing and counselling (PITC) virtually eliminated (4).

Despite introducing more targeted HIV testing approaches such partner testing services in Uganda, the overall HTS yield remained relatively constant from 3.1% in 2017, peaking at 3.8% in 2018 and regressing to 3.1% in 2019. Whereas index testing (including partner testing services) provided significant yield (20%) in 2019, the overall contribution to case identification by this HTS modality was only 15.4%, with provider-initiated counselling and testing other (PITC-other) modality contributing 53% of all HIV-positive cases identified in 2019 (5). Large facility-based entry points like PITC, however, reach large numbers of clients and identify more PLHIV in absolute numbers than targeted strategies, even if they are of lower yield. As a result, in recent years, the vast majority of newly identified PLHIV have been identified through facility-based testing (6). For example in 2018, PITC at outpatient departments (OPD) and facility-based voluntary counselling and testing (VCT) accounted for approximately 68% of all newly diagnosed PLHIV (7).

Given resource constraints, ministries of health are looking for opportunities to improve HIV testing efficiency through reducing the total number of people tested for HIV while targeting to identify the same or higher numbers of PLHIV hence increasing the positivity rates. To achieve this, many countries are institutionalized HIV risk screening tools to identify high risk populations. These tools use a combination of clinical and behavior-based criteria to identify individuals with a higher risk of HIV infection who are then prioritized for testing. Screening tools have been used successfully for pediatric and adolescent clients to identify children/adolescents living with HIV and are used to screen-in children and adolescents for testing using socio-demographic and clinical variables (8). However, evidence around use of screening tools in adults is limited (9). Evaluations of behavior-based risk algorithms in the United States and Malawi, to identify sexually transmitted infection (STI) indicate the tools have varying sensitivities and specificities (10). Many countries have adopted the use of risk-based screening tools among adult populations, which screen out significant numbers of people and only test those defined as eligible. The underlying hypothesis is to reduce the total number of tests while increasing the positivity rate.

The HTS program at the Ministry of Health (MOH) in Uganda introduced an HIV risk screening tool to determine eligibility for HIV testing among clients attending OPD clinics. Beginning in October 2019, the MOH deployed this tool for use in 24 health facilities, targeting clients aged 15 years and above in both OPD and VCT departments.

The screening tool included following questions:

- Does patient have co-morbidities or an exposure risk?
  - Presumptive TB
  - New perpetuators and survivors of SGBV
  - A reactive HIV self-test
  - Elicited through index testing
  - Accidentally exposed to HIV
  - Diagnostic HTS (unconscious, critically ill, mentally impaired)
- Has the client had an HIV test in the last 12 months?
- Has the client tested within the last 3 months? If No does the client meet any of the following criteria:
  - Client has had unprotected sex with partner(s) of unknown HIV status or known HIV positive status since the last negative HIV test?
  - HIV negative partner(s) in a discordant relationship and has not had an HIV test within the past 3 months
  - Client has diagnosis of sexually transmitted infection (including Hepatitis B) after previous negative HIV test
  - Client with TB, STI, Hepatitis B, symptomatic of HIV, or is on PEP and Tested HIV Negative at least 1 month ago

A client was eligible for HIV testing if any of the responses was “Yes” and ineligible for testing if all the responses were “No”.

We aimed at determining the operational performance of the screening tool in public health facility settings, by assessing the diagnostic characteristics of the tool in terms of sensitivity, specificity, predictive values, and number needed to test to identify an individual with HIV. We also aimed to determine the cost implication of using or not using the screening tool by analyzing the estimated comparative costs of testing with and without screening as a secondary outcome.

## Materials and methods

### Design

We conducted a retrospective secondary data analysis of de-identified and anonymized HIV testing program data, collected and reported by 24 health facilities in Uganda. The de-identified data was requested from the MOH for secondary analysis and upon approval, was extracted from the DHIS2 by district biostatisticians. Data analysis was performed between March and April 2020 on all HIV risk screening data collected and reported between October and November 2019. The primary outcome was to compare the sensitivity and specificity of the screening tool with the standard of care (testing without screening), and the secondary outcome was to estimate the cost savings on using the screening tool compared to the standard of care.

### Settings

Ministry of health Uganda deployed the adolescent and adult HIV risk screening tool to determine eligibility for HIV testing among clients attending OPD clinics in 24 health facilities in October 2019. The screening tool was deployed at various levels of the health care system, including primary, secondary, and tertiary health facilities. The use of the screening tool was integrated into routine care, and all clients who sought HTS in the OPD and VCT clinics were consented and screened by health care providers. Health workers used the risk screening tool to determine clients’ eligibility for HIV testing. Because clients who attended HIV testing points in the OPD and VCT had turned up for HIV testing, the use of the screening did not exclude anyone from testing, hence both clients who were categorized as eligible or ineligible upon screening were offered HIV testing services including testing and linkage to posttest services. In line with national guidelines, the screening tool was only administered to clients over the age of 15 years. Screening and testing information was recorded using the national health management information system using primary data collection registers. The information was decoded, anonymized, and reported by the health facilities in the district health information system (DHIS2).

### Study participants

The data analysis included entries of all clients aged 15 years and above, who were screened for HIV testing eligibility at the 24 health facilities in the months of October and November 2019.

### Study variables

The predictor for the primary outcome was binary, “eligible for screening”, or “Not eligible for screening”. And the outcome was the HIV test result “Tested HIV Positive”, or “Tested HIV Negative”. These were stratified by social demographic characteristics including age, gender, and marital status. From the eligibility and the HIV testing results variables, we computed the predictive values Sensitivity, specificity, and number needed to test (NNT) to identify one HIV positive person with or without screening. For the secondary outcome, we estimated the unit cost needed to identify one HIV positive client with or without risk screening from which we determined the cost difference.

### Sample size and data sources

We included all (census) data submitted into the DHIS2 for clients screened for HIV eligibility at the 24 health facilities in the two months’ period (October and November 2019). The data source was DHIS2 (secondary data) from districts where the health facilities belonged. By considering all the data submitted for all facilities, we excluded selection bias which would result from using a small sample. The choice of the 24 health facilities was because, these were the first facilities to use the screening tool and data from these facilities would inform further scale up.

### Data analysis

Data was abstracted from the DHIS2 by district biostatisticians and shared with the authors for analysis. The data was checked for consistency and accuracy, was cleaned using excel software, and exported into STATE/SE 15 (StataCorp, College Station, Texas) software for analysis. We included a record which reported both the screening eligibility and a documented HIV test result. We excluded entries with missing screening eligibility, missing HIV test results, and those reporting an age below 15 years. HIV positivity rates (also computed statistically as PPV) with and without screening were calculated allowing for clustering by facility. Sensitivity, specificity, and number needed to test (NNT) were also calculated. Logistic regression with random effects was used to estimate the odds ratio (OR) associated with HIV status for each variable.

Cost estimates were calculated based on commodity and human resource (HR) required to conduct HIV testing for clients who were categorized as eligible. Commodity costs were based on public procurement cost of HIV rapid diagnostic tests in Uganda at the time of screening. Human resource costs were calculated using the average salary for counselors in public health facilities. Time requirements for standard of care (no screening in HTS) assumed the time for counseling and testing as was outlined in national guidelines while screening time assumptions were based on implementing partner reports. Estimated costs did not include operational costs, such as training and printing of risk screening tools.

### Ethical considerations

The authors analyzed anonymized retrospective secondary program data which did not include client identification information. Neither the authors nor the district biostatisticians interacted with any primary client records before, during or after the data analysis. The analyzed data had been decoded at the time of reporting into the DHIS2 hence access to this data did not pose breach of confidentiality or would not potentially lead to identification of clients. For this reason, the authors did not seek IRB approval but sought clearance from the Ministry of Health to access the secondary data from district DHIS2 systems through district biostatisticians.

## Results

A total of 19,854 clients were screened for HIV testing eligibility, we excluded 137 records due to missing HIV testing information, and an additional 13 records were excluded because the documented age was below <15 years (Figure 1).

**Figure 1:**
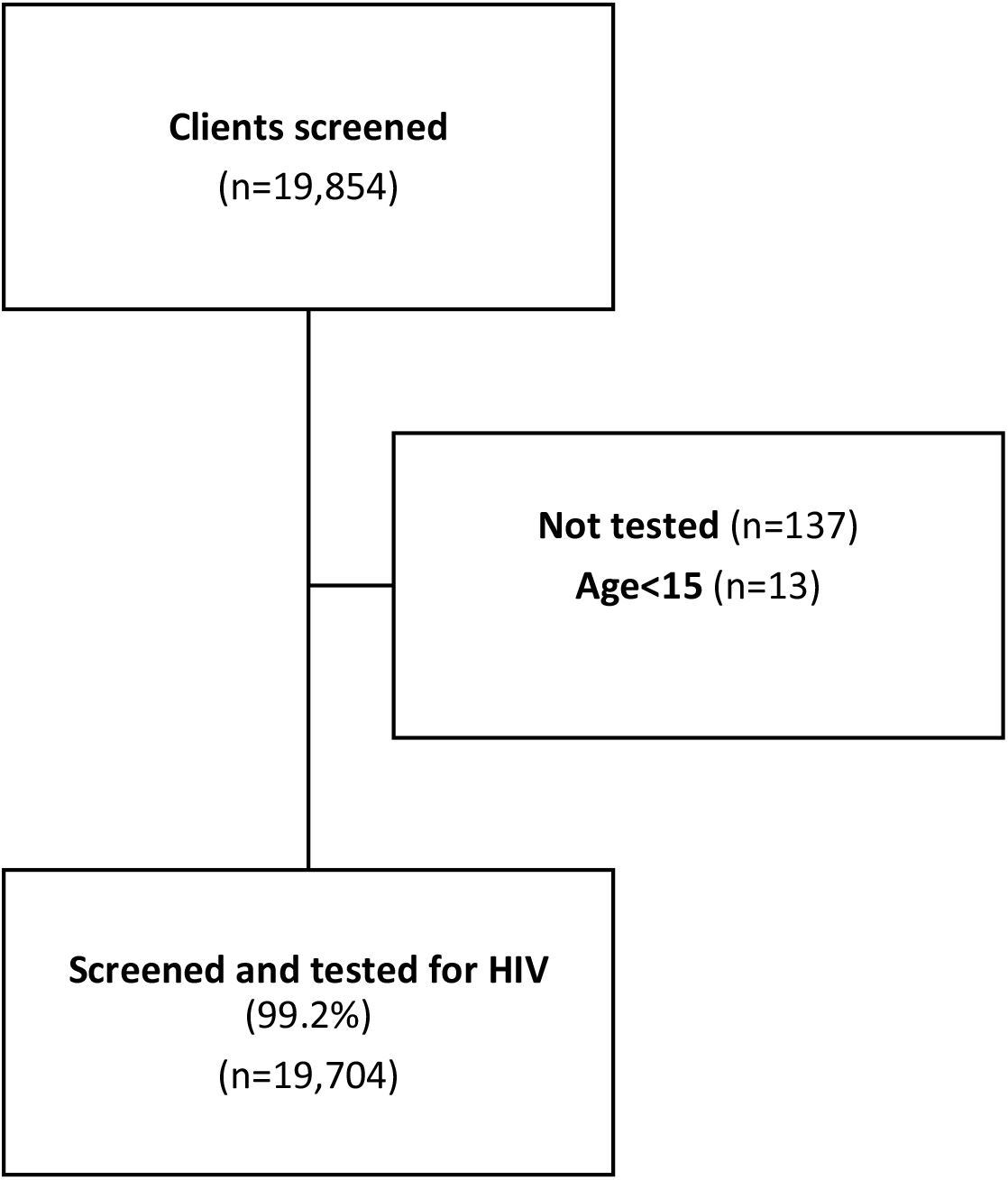
Study flow chart.

Of the remaining 19,704 (99.2%) clients, 12,971 (66%) were female, with a median age of 27 yeas (IQR: 21-35). (Table 1).

**Table 1:** Baseline characteristics stratified by HIV status.

| Characteristics | All<br>N=19,704 | HIV Status |  | Univariate OR<br>(95% CI) |
| --- | --- | --- | --- | --- |
|  |  | Positive<br>n= 732 (%<sub>col</sub>) | Negative<br>n= 18,972(%<sub>col</sub>) |  |
| Age (years) |  |  |  |  |
| 15-19 | 3,379 (17%) | 42 (6%) | 3,337 (17%) | 1 |
| 20-35 | 11,469 (58%) | 457 (62%) | 11,012 (58%) | 3.10 (2.08-4.63) |
| 36-50 | 3,310 (17%) | 171 (24%) | 3,139 (17%) | 4.21 (2.78-6.35) |
| 50+ | 1,546 (8%) | 62 (9%) | 1,484 (8%) | 3.33 (2.00-5.56) |
| **Median age (IQR)** | 27 (21-35) | 30 (25-39) | 26 (21-35) |  |
| **Gender** |  |  |  |  |
| Female | 12,971 (66%) | 475 (65%) | 12,496 (66%) | 1 |
| Male | 6,733 (34%) | 257 (35%) | 6,476 (34%) | 1.01 (0.83-1.24) |
| **Marital Status** |  |  |  |  |
| Married | 12,071 (61%) | 464 (63%) | 11,607 (61%) | 1 |
| Divorce/ Separated | 1,544 (8%) | 115 (16%) | 1,429 (8%) | 2.11 (1.59-2.81) |
| Single | 5,628 (29%) | 128 (18%) | 5,500 (29%) | 0.60 (0.43-0.85) |
| Widowed | 461 (2%) | 25 (3%) | 436 (2%) | 1.60 (0.88-2.88) |
| **Screening tool**<br>(Yes, to any eligibility criteria) | 14,879 (76%) | 664 (91%) | 14,215 (75%) | 3.60 (2.30-5.62) |

The overall positivity rate (cluster weighted prevalence of HIV) was estimated at 3.71% (95% CI: 3.06-4.50), which would be the yield without screening. Among those screened, the positivity rate was 4.5% (95% CI: 4.1%-4.8%). The sensitivity of the tool was 90.7% (95% CI: 88.4%, 92.7%) while the specificity was 98.6% (95% CI: 98.2%-98.9%). With screening, the number needed to test (NTT) to identify one PLHIV reduced from 27 to 22. See Table 2.

**Table 2:**
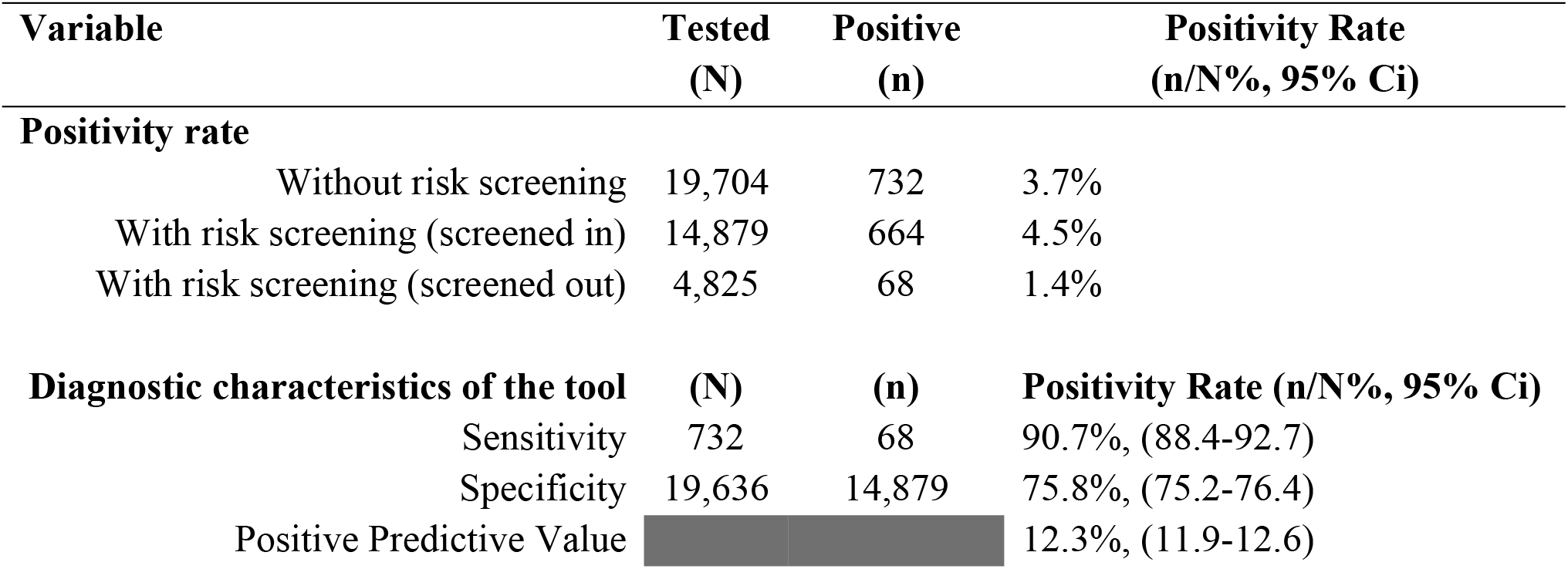
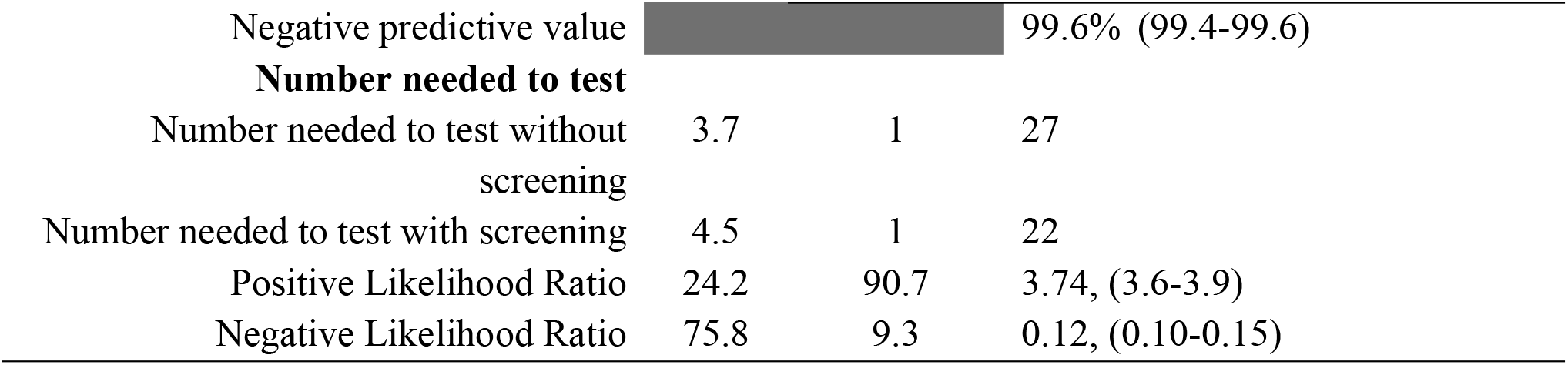
Diagnostic characteristics of the adult HIV risk screening tool

Overall screening for HIV testing eligibility using the screening tool would have led to 24.5% (4,825/19,704) reduction in testing volume but 9.3% (68/732) of PLHIV would have been missed as they were misclassified as not eligible for testing. The cost per PLHIV identified fell by 3% from $69 without screening to $66.9 with implementation of the screening tool (Table 3).

**Table 3:** Cost analysis for implementing HIV screening using a risk screening tool at an outpatient department (OPD) in Uganda.

| **HR and commodity costs for current standard of care compared with screening in OPD** |  |  |  |
| --- | --- | --- | --- |
|  | **Standard of Care** | **Screening in OPD** | **Screening tool savings** |
| **Total number of tests (A1)** | **19,704** | **16,764** | 2,951 |
| **Total cost** | **\$44,357** | **\$40,222** | **\$4,138** |
| Commodities | \$25,825 | \$22,036 | \$3,790 |
| Human resources | \$18,532 | \$18,184 | \$348 |
| **Cost per PLHIV identified** | **\$69.05** | **\$66.91** | **\$2.14** |
| Commodity cost per PLHIV identified | \$40.2 | \$34.4 | \$5.9 |
| Human resources per PLHIV identified | \$28.85 | \$28.31 | \$0.54 |

## Discussion

Over the past years, there has been a strong narrative supporting the use of risk screening tools to improve testing efficiency with very limited evidence on their impact. This program data analysis identifies operational gaps in HIV case identification among clients who seek health services at outpatient departments and highlights how HIV risk screening tools may misclassify HIV positive clients as “not at risk” of being HIV positive. These findings relate to those of Antelman and colleagues (11), where a risk screening tool for children and adolescents in Tanzania was reported to miss an unacceptably high proportion (36%) of HIV-positive children. Such missed opportunities may propagate HIV transmission resulting from being unaware of the positive HIV status and may lead to delayed diagnosis and linkage to treatment resulting into AIDS related deaths.

Whereas secondary analysis of routine programmatic data from Uganda shows the screening tool could reduce testing volumes by 24% hence apparently saving on the cost per HIV positive case identified, screening resulted in a marginal increase in positivity rate from 3.71% to 4.5%. Of more concern is the number of clients who were misclassified as ineligible for HIV testing, yet they were HIV positive. Health workers in Uganda routinely provide HIV testing demand generation health talks (information giving) at the OPD and VCT waiting areas, which include information on how to opt in for HIV testing. By opting to test for HIV, it means these clients have a perceived risk of being HIV positive and would not need to undergo another layer of screening; thus, without screening, all clients who choose to test for HIV would be tested.

Considering the risk screening tool’s inefficiency on a national scale, the number of PLHIV that would be missed through screening at a 90.7% sensitivity is significant. In 2021 for example, Uganda conducted approximately 4,608,652 HIV tests of which 1,753,704 (38%) tests were in OPD. Applying the screening tool to that population would result in missing 5,002 PLHIV, hence it is essential for Ministry to weigh this impact as it scales up implementation.

From a cost perspective, screening did slightly reduce the cost per PLHIV identified by about 3% and did result in overall program savings from the lower commodity requirements for reduced testing volumes. However, these savings only accounted for human resource and commodity costs and do not reflect the full costs of implementing screening tools which would include training, printing, dissemination of tools, monitoring, and evaluation among others. In addition, these savings were at the expense of missed PLHIV and would be offset by the cost to reach these missed PLHIV through alternative strategies such as index testing. Facility-based testing is generally less expensive compared to community testing strategies such as outreach, and mobile testing hence all opportunities need to be maximized to identify HIV positive clients who present at the health facilities. If clients living with HIV are screened out at facilities, ministries need to consider if they will ultimately be able to identify these clients through alternative, more expensive testing models. Moreover, it is of ethical concern that HIV positive clients are classified as HIV negative.

Routine use of risk screening tools would require training and supervision of HIV testers to minimize user errors that lead to misclassification of clients. During monitoring and supervision visits by ministry of health, it was established that some healthcare workers did not follow the screening standard operating procedure leading to misclassification of clients. Although training and ongoing mentorship would improve healthcare worker capacity to screen, this would probably only address the misclassification bias to a limited extent; for example, if clients do not feel comfortable answering the screening questions truthfully, the risk screening tool would not detect the misclassification. Improving sensitivity of the tool would require formulation of less stigmatizing questions and more private spaces for responses. This is an area for further study to determine the extent to which misclassification bias can be reduced.

The costs and implications of failing to identify PLHIV within health facilities where they could be identified and linked to care, may quickly outweigh savings in testing commodities and calls for strategic reforms by countries to consider alternatives to risk screening tools. Of recent, there is growing advocacy for countries to adopt HIV self-testing (HIVST) as a screening approach for clients seeking HIV testing at both facility and community (test for triage). Scaling up HIVST would require formulation or adoption of HIVST policies and a considerable financial investment to roll out these policies, plus commodity management. Recent research in Malawi has shown the potential of HIVST to expand testing coverage, while reducing HR time and limiting the risk of screening PLHIV out, given the much higher sensitivity of antibody screening platforms compared to risk-based screening tools (12).

Much as countries are exploring use of HIV risk screening tools to identify PLHIV more efficiently and make better use of available resources, the evidence presented above clearly illustrates the tradeoffs involved in implementing these tools. Lowering HIV testing volumes comes at the expense of screening out PLHIV who have presented for testing in facilities. Furthermore, any overall savings made through use of risk screening tools are significantly offset by the added human resource costs during program implementation. Given that majority of PLHIV have been identified globally and narrowing this to individual countries, the reliance on risk screening tools to classify who is likely to be HIV positive may be counterproductive especially in low HIV prevalence countries or in high prevalence countries but with a low HIV treatment adjusted prevalence. For such countries, an antibody/antigen screening test would be most ideal.

## Limitations

This was a secondary analysis of routine program data from 24 health facilities that were not randomly selected and may not represent over 3,000 health facilities that offer HTS in the country. Costing included commodity and human resource costs, as these are primary cost drivers for HTS, but was not exhaustive and did not include operational costs. HR time requirements were estimated based on guidelines and implementing partner reports, rather than time-in-motion studies.

## Conclusion

The use of HIV risk screening tool in OPD settings in Uganda demonstrated improved HIV testing efficiency by reducing testing volumes but resulted in screening out of a significant number of people living with HIV. There was minimal cost savings earned through testing fewer people, but these would be offset by the cost to reach the missed PLHIV through alternative and more expensive HIV testing strategies such as index testing.

## Recommendations

The team recommends use of scientifically validated HIV risk screening tools by countries; ministries should provide regular support supervision and mentorship to all HIV testers to ensure adherence to the risk screening SOPs, and to limit misclassification of clients seeking HTS, facility based HIVST (HIV antibody test) should be adopted. Scientific validation of risk screening tool using a statistically representative sample is recommended to generate generalizable results.

## Data Availability

The data underlying the results presented in the study are available from Ministry of Health Uganda, STD/AIDS Control Program upon request via

## Acknowledgements

Special thanks to the staff of Uganda Ministry of Health, STD/AIDS Control program, Health workers at the 24 healh facilities where secondary data was abstracted, PEPFAR-CDC Uganda and CHAI Uganda.

